# Epidemiology of diabetic foot in Bukavu

**DOI:** 10.64898/2026.06.27.26356745

**Authors:** Jérôme Mbala Ethienne, Daniel Kakusu, Mapenzi Ndagonywa Philémon

## Abstract

The prevalence of diabetes is increasing in all countries, and diabetic foot is a complication with serious functional consequences. Due to its prevalence and the morbidity it causes, diabetic foot has become a public health problem. Our retrospective cross-sectional study is primarily descriptive. Its aim is to determine the frequency of diabetic foot in the surgical department of the Panzi General Referral Hospital during the period from January 2017 to December 2021. During these five years, the hospital recorded 745 hospitalized patients, of whom 25 were included in this study. The frequency was 3.4%, the mean age was 46.84 ± 22.4 years (range 3 to 80 years), and patients aged 41 to 60 years were the most affected, representing 44% of our series. There was a female predominance, with 68% of the cases studied, and a sex ratio of 0.47. The majority of patients came from urban areas (64% of cases). Arteriopathy was the most frequent type of diabetic foot (48%), with gangrenous lesions accounting for another 48%, and type 2 diabetes being the most prevalent (68%). Conversely, 94% of patients admitted to the surgical department of the Panzi General Referral Hospital were discharged with their lesions stabilized. Diabetic foot is common among diabetic patients in South Kivu. Combating this scurge requires patient and healthcare staff education, as well as multidisciplinary and coordinated care. Our study and literature review highlighted the epidemiology of diabetic foot in order to determine its prevalence in South Kivu.

## Introduction

Diabetic foot encompasses any infection, ulceration, or destruction of the deep tissues of the foot associated with neuropathy and/or peripheral arterial disease of the lower limbs in diabetic patients (1). It constitutes a public health problem. According to the WHO report in 2011, micro- and macrovascular complications are a major cause of morbidity and mortality worldwide. Among these, neuropathy and peripheral arterial disease of the lower limbs are two cardinal features of diabetic foot; their respective prevalence is estimated at 40% in the diabetic population and 50% in cases of ulceration (2).

The frequency and severity of foot problems in people with diabetes vary from region to region, with an annual incidence of around 2 to 4% in developed countries, and probably even higher in low-income countries with a prevalence of around 19% to 34% (3). It is a frequent and serious complication of diabetes, with a very high rate of lower limb amputation and often dramatic socio-economic and psychological consequences for the population. (1).

In 2005, according to the national health insurance fund for salaried workers, the prevalence of treated diabetic foot in the metropolitan population was 3.8% (3).

China in 2018 provided the most striking illustration of the progression of diabetes in major emerging countries, with a diabetic population (diagnosed and undiagnosed) of 114 million individuals, the largest in the world, with type 2 diabetes constituting the vast majority of cases (4). The most representative survey conducted in 2013 revealed that the rate of diabetic foot (detected and undetected) was 10.9%, a percentage close to that of the United States in 2011-2012 (5). In France, a nationally representative sample study of people with diabetes reports a prevalence of foot ulcers, healed or unhealed, of 6% according to patient data and 1.5% according to physicians (6). The annual incidence of foot ulcers in people with diabetes is estimated to be between 0.5% and 3%. Overall, 15% of people with diabetes will experience a foot ulcer during their lifetime (6) . In Africa, foot lesions in people with diabetes are unfortunately very common. They account for 15% to 25% of hospitalizations. Often, poverty, poor hygiene, and walking barefoot interact to worsen the impact of foot lesions caused by diabetes (7). In 2011 in sub-Saharan Africa, diabetic feet were estimated at 5.7 million, with 737,090 cases in the Democratic Republic of Congo (DR Congo) for the same year (8). In Algeria, the number of consultations for foot in diabetic people in emergency or outpatient follow-up is 2626 patients, or 219 patients per month over the last year (June 2018-June 2019) (9). In Togo in 2015, the prevalence of diabetic foot was 12.90%. The starting point for foot lesions was trauma resulting in a superinfected wound in 70.97% of cases (10) . In the Democratic Republic of Congo (DRC), we found no studies concerning this problem. We were unable to calculate the incidence or prevalence due to the lack of reliable regional or national statistics on diabetic foot. The province of South Kivu, including the city of Bukavu, is clearly affected by this situation, with many undiagnosed cases, making it difficult for healthcare workers to establish realistic statistics regarding this pandemic.

Eu In light of all the above, we asked ourselves the following question:

What is the frequency of diabetic foot in the province of South Kivu? Given the research question, this work will have the following hypotheses:

- The frequency of diabetic foot is said to be high at HGR/PANZI.
- Women and the elderly will be more affected.
- Hypertension would be the associated risk factor.

The general objective of this study will be to determine the frequency of diabetic foot at the PANZI General Referral Hospital.

Specifically:

- Determining the frequency of diabetic foot in South Kivu
- Identify the most affected age group and gender
- Identify risk factors associated with this condition

The value of this study lies in the subsequent use of its results by a multidisciplinary team (obstetricians, anesthesiologists, neonatologists, cardiologists and neurologists, psychologists and even sociologists) to improve the management of this pathology, while placing particular emphasis on its complications and severity in order to reduce maternal and fetal morbidity and mortality.

### Materials and methods Nature and Period of Study

The present study is a retrospective cross-sectional study with an essentially descriptive aim was conducted by analyzing the medical records of 745 patients in our structure, from 1st January, 2017 to 31st December, 2021, covering the period of our study.

### Study Framework

The records of the surgical department of the Panzi general referral hospital allowed us to conduct our study.

### Study population

This work mainly concerns diabetic patients who came for consultation, were hospitalized for infection, ulceration or destruction of deep tissues of the foot throughout our study period.

### Sample Inclusion criteria

We included all diabetic patients who came for consultation or were hospitalized for infection, ulceration, or destruction of deep tissues of the foot throughout our study period.

### Exclusion criteria

We exclude from our work all patients who came for consultations or were hospitalized for surgery for other health problems.

### Data Collection Variables of interest

The variables of interest were: age, sex, origin, marital status; occupation, religion, medical history, clinical data, evolution.

### Data Collection Instrument

The survey questionnaires allowed us to collect the data.

### Data Analysis

The data will be presented as percentages in various tables. This will allow us to properly interpret the results of our work (Study). To interpret our results, we used statistical calculations and information including:

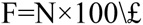

We used EPI INFO 7 software, Excel and Word for text entry, to facilitate the task of encoding data and to perform some calculations.

### Ethical considerations

Our study respected the freedom and consent of the respondents. Anonymity was also observed throughout the data collection process.

## Results

### Prevalence

**Table I.**
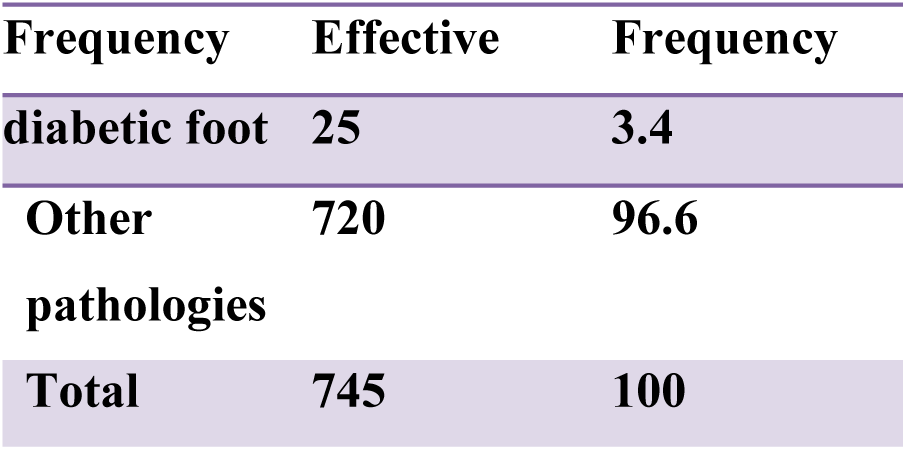
Frequency of diabetic foot.

This table shows that the frequency of diabetic foot in the surgical department of Panzi Hospital was 3.4%.

#### Socio-demographic data

**Table II.**
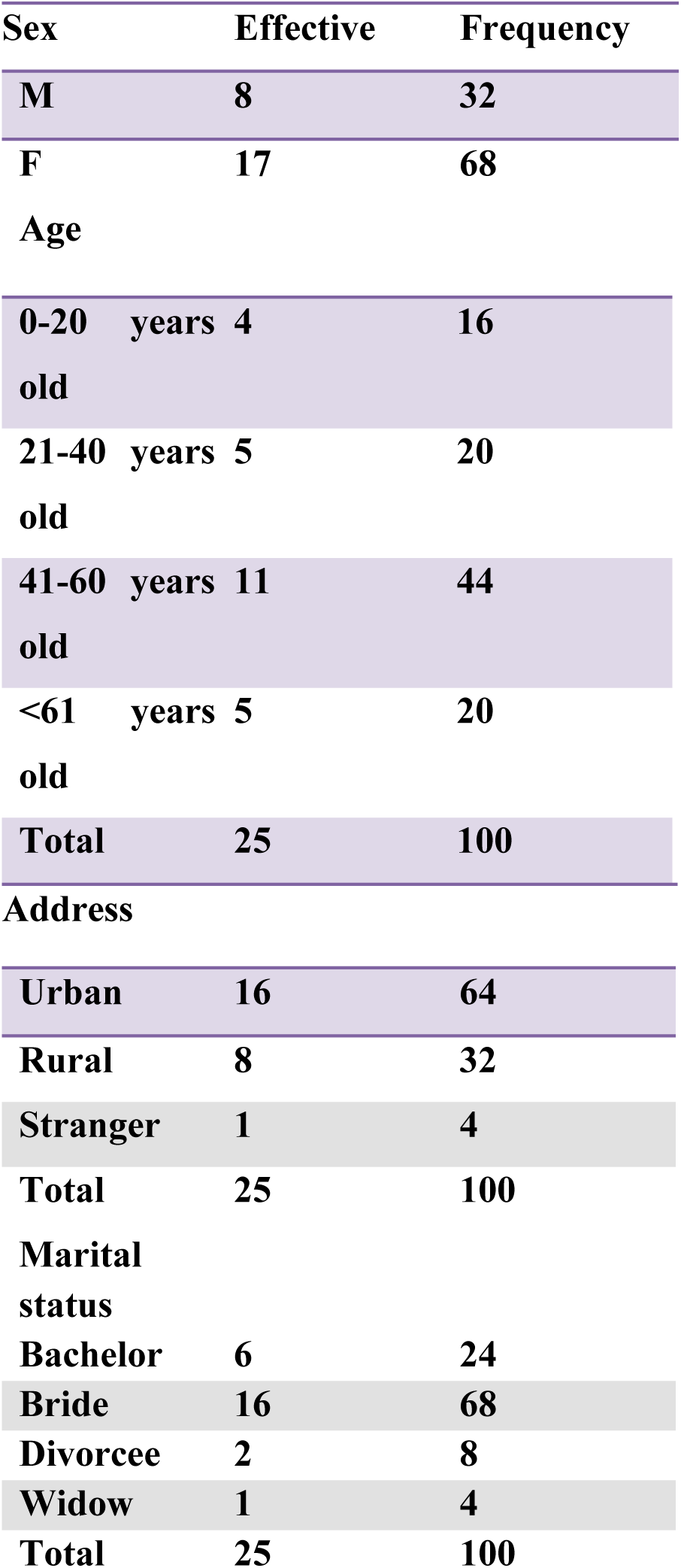
Distribution of patients according to socio-demographic data:

| Sex | Effective | Frequency |
| --- | --- | --- |
| M | 8 | 32 |
| F | 17 | 68 |
| Age |  |  |
| 0-20 years old | 4 | 16 |
| 21-40 years old | 5 | 20 |
| 41-60 years old | 11 | 44 |
| <61 years old | 5 | 20 |
| Total | 25 | 100 |
| Address |  |  |
| Urban | 16 | 64 |
| Rural | 8 | 32 |
| Stranger | 1 | 4 |
| Total | 25 | 100 |
| Marital status |  |  |
| Bachelor | 6 | 24 |
| Bride | 16 | 68 |
| Divorcee | 2 | 8 |
| Widow | 1 | 4 |
| Total | 25 | 100 |

The mean age was 46.84± Standard deviation is about 22.445 with extremes of 3 years and 80 years and patients aged over 35 years were the most affected, representing 76% of our series.

There was a female predominance, with 68% of the cases studied representing a sex ratio of 0.47. Furthermore, the majority of patients came from urban areas (64%). Married patients accounted for 68% of cases.

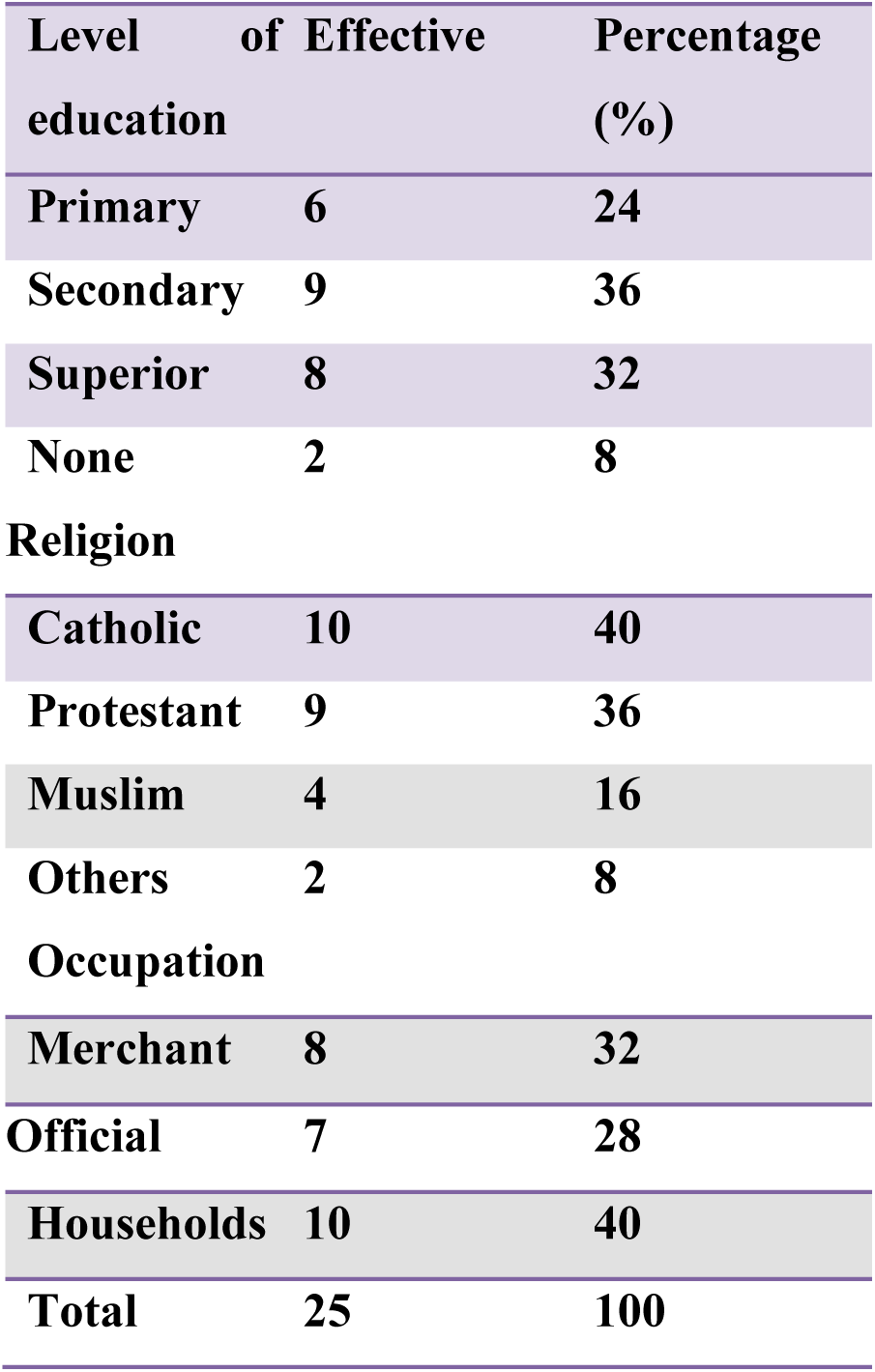

Regarding religious affiliation, Catholics are the most affected by foot ulcers (40%), followed by Protestants (36%). Patients with a humanities degree are more likely to be affected (36%), while those with a primary school education are more likely to be affected (24%).

Housewives represented 40% of patients, followed by shopkeepers with 32%.

## Background

### Family history

**Table III.**
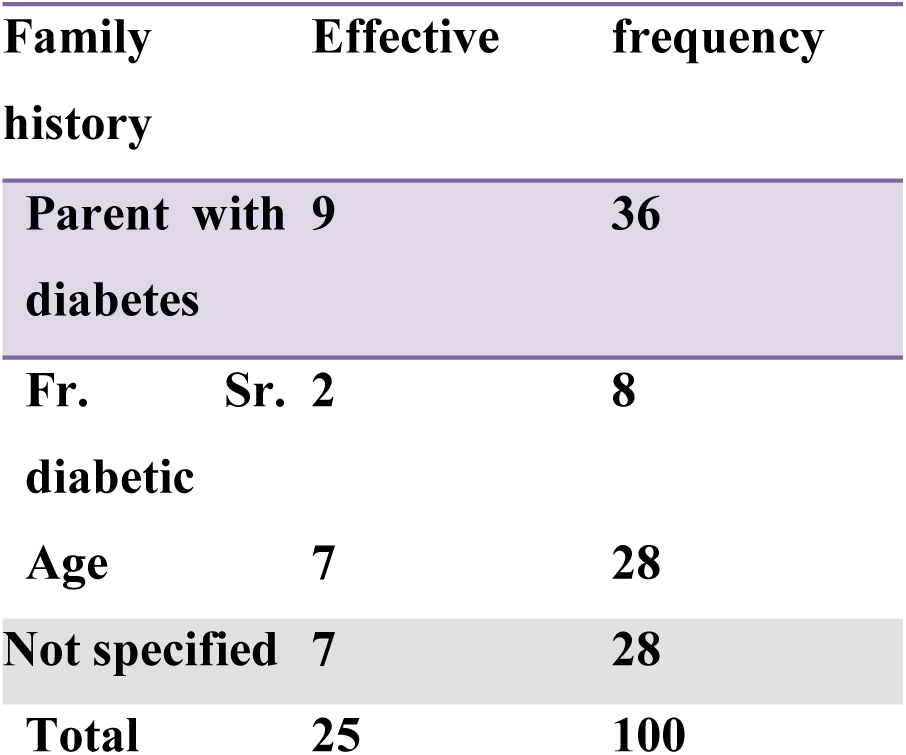
Distribution of patients according to their family history:

In our series of studies, at least 36% of parents were diabetic.

### Medical history

**Table IV.** Distribution of patients according to their medical history.

| Medical history | Effective | frequency |
| --- | --- | --- |
| HTA | 9 | 36 |
| Obesity | 5 | 20 |
| Dyslipidemia | 4 | 16 |
| Smoking | 5 | 20 |
| Not specified | 2 | 8 |
| Total | 25 | 100 |

Hypertension (HTA) accounts for 36% and obesity for 20% of the conditions that lead to diabetes and PD.

### Clinical data

#### The type of injury

**Table V.**
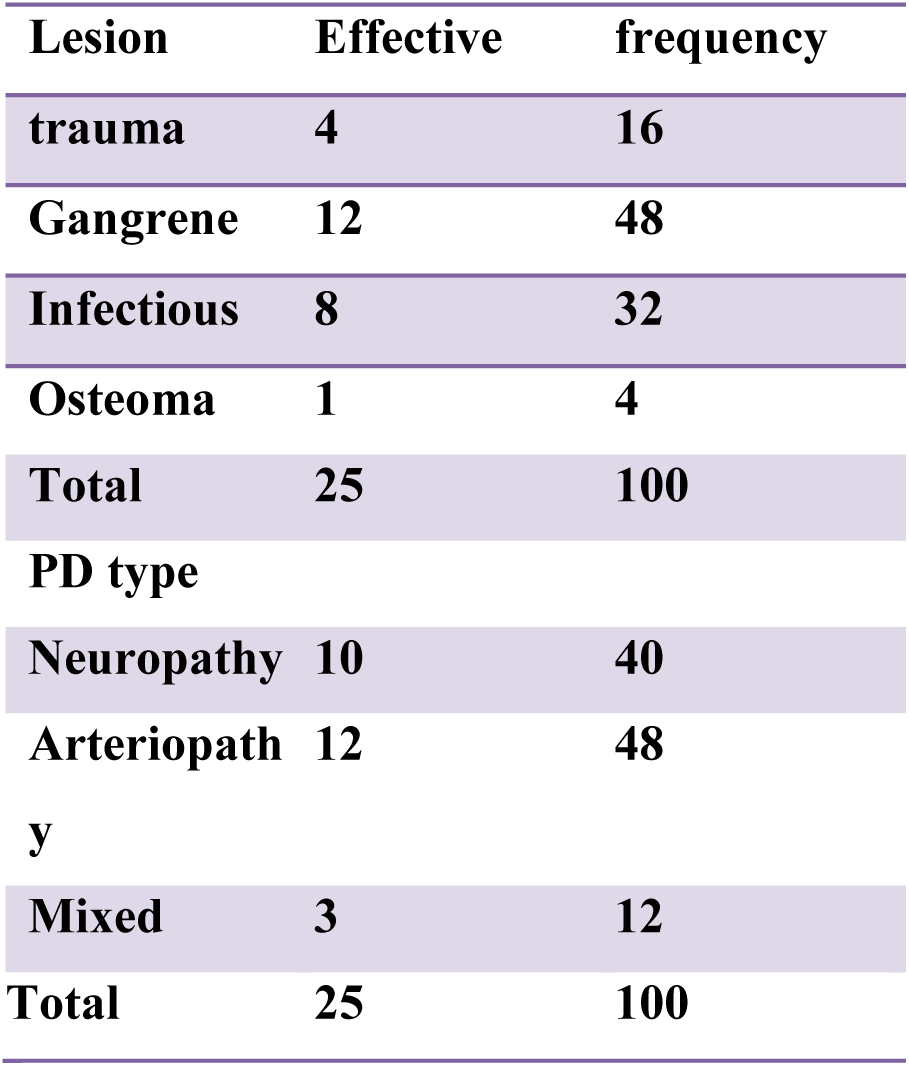
Distribution of patients according to the type of lesion.

Gangrenous lesions represent 48% of the cases studied in our study series and the most frequent type of diabetic foot is arteriopathy with 48%.

### The duration of the injury

**Table VI.** Distribution of patients according to the duration of the lesion.

| Duration | Effective | frequency |
| --- | --- | --- |
| 0 to 30 days | 9 | 36 |
| 31 to 60 days | 14 | 56 |
| Ignore | 2 | 8 |
| Total | 25 | 100 |

This table shows that 36% of diabetic patients consulted a doctor within 30 days. The average duration of the lesion is 30 days, with a range from 30 to 60 days.

### Type of diabetes

**Table VII.** Distribution of patients according to type of diabetes.

| Type of diabetes | Effective | frequency |
| --- | --- | --- |
| Type 1 | 8 | 32 |
| Type 2 | 17 | 68 |
| Total | 25 | 100 |

Type 2 diabetes is the most frequent cause of foot ulcers in diabetic individuals, with a frequency of 68%.

### Patient progress

#### Evolution

**Table IX.**
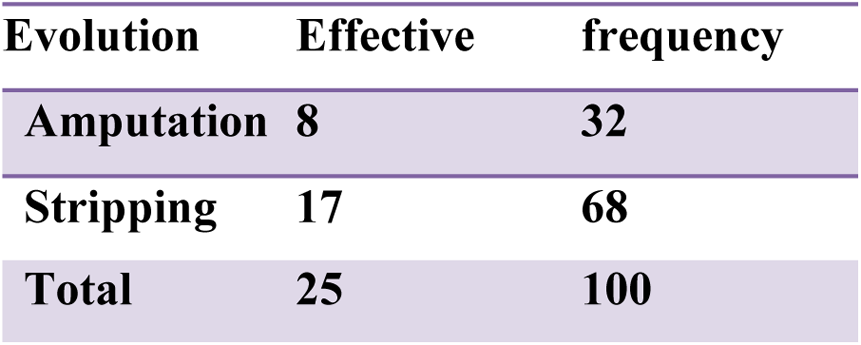
Distribution of patients according to the progression:

| Evolution | Effective | frequency |
| --- | --- | --- |
| Amputation | 8 | 32 |
| Stripping | 17 | 68 |
| Total | 25 | 100 |

In most patients treated for diabetic foot, their condition progresses to amputation.

### Exit method

**Table X.**
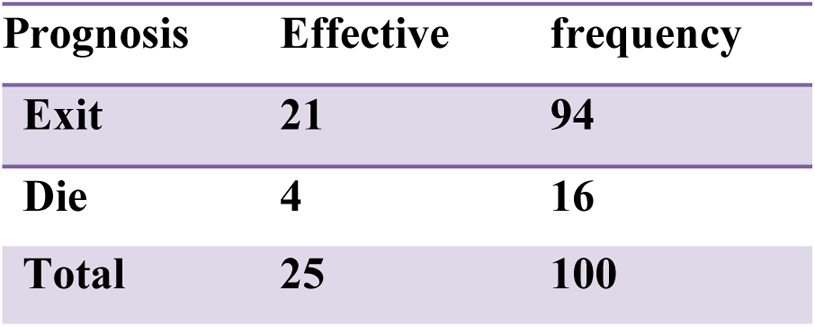
Distribution of patients according to discharge method.

This prognosis table shows that 94% of patients admitted to the surgery department of the PANZI General Referral Hospital were discharged with improved injuries and 16% lost their lives.

### Linking variables

#### Medical history and types of injuries

**Table X.**
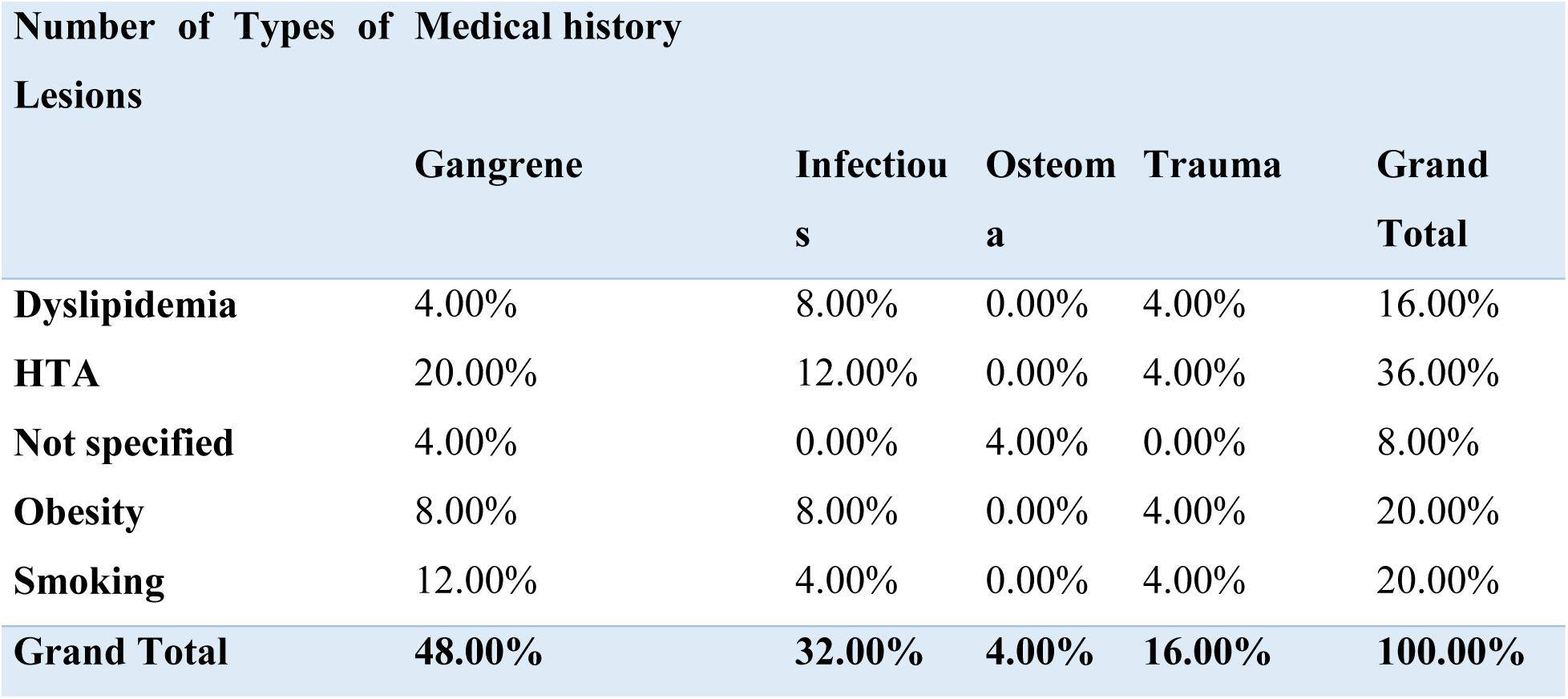
Distribution of patients according to lesion type based on medical history.

| Number of Types of Medical history Lesions |  |  |  |  |  |
| --- | --- | --- | --- | --- | --- |
|  | Gangrene | Infectious | Osteoma | Trauma | Grand Total |
| Dyslipidemia | 4.00% | 8.00% | 0.00% | 4.00% | 16.00% |
| HTA | 20.00% | 12.00% | 0.00% | 4.00% | 36.00% |
| Not specified | 4.00% | 0.00% | 4.00% | 0.00% | 8.00% |
| Obesity | 8.00% | 8.00% | 0.00% | 4.00% | 20.00% |
| Smoking | 12.00% | 4.00% | 0.00% | 4.00% | 20.00% |
| Grand Total | 48.00% | 32.00% | 4.00% | 16.00% | 100.00% |

This cross-table illustrates the impact of medical history on the types of lesions, explaining that 36% of patients who had hypertension, 20% of them had gangrene, 12% had infectious lesions and 4% had traumatic lesions.

### Type of diabetic foot in relation to patient address

**Table XI.** Distribution of patients by type of diabetic foot and their addresses.

| Number of PD Types | Address |  |  | Grand Total |
| --- | --- | --- | --- | --- |
|  | Stranger | rural | urban |  |
| Arteriopathy | 4.00% | 8.00% | 36.00% | 48.00% |
| Mixed | 0.00% | 4.00% | 8.00% | 12.00% |
| Neuropathy | 0.00% | 20.00% | 20.00% | 40.00% |
| Grand Total | 4.00% | 32.00% | 64.00% | 100.00% |

In this table, among the 48% of patients with arteriopathy, 36% were of urban origin, 8% of rural origin and 4% were foreigners.

For neuropathy, 32% of the patients came from rural areas (8 patients) and 20% of them presented with neuropathy as a type of diabetic foot.

### Type of diabetes and its progression

**Table XII.** Distribution of patients by type of diabetes and their progression.

| Number of evolutions | Types of diabetes |  |  |
| --- | --- | --- | --- |
|  | Amputation | stripping | Grand Total |
| Type I | 8.00% | 24.00% | 32.00% |
| Type II | 36.00% | 32.00% | 68.00% |
| Grand Total | 44.00% | 56.00% | 100.00% |

In this table, among the 17 type II diabetic patients, only 36% underwent amputation and 32% were treated with debridement. Among those with type I diabetes (32%), only 8% underwent amputation.

### Background and discharge method

**Table XIII.** Distribution of patients according to their medical history and discharge methods.

| Exit method |  |  |  |  |  |  |
| --- | --- | --- | --- | --- | --- | --- |
| Number of background | Dyslipidemia | HTA | NP | Obesity | Smoking | Total |
| Death | 4.00% | 4.00% | 0.00% | 4.00% | 4.00% | 16.00% |
| Exit | 12.00% | 32.00% | 8.00% | 16.00% | 16.00% | 84.00% |
| Total | 16.00% | 36.00% | 8.00% | 20.00% | 20.00% | 100.00% |

This table illustrates that among the 84% of patients who were successfully discharged from the hospital, 12% had hypertension, while chronic smokers and obese individuals shared the same percentage at 16%. 8% of them had an unspecified medical history.

## Discussion of results

### Prevalence

During our study, 25 patients out of a total of 745 were hospitalized for diabetic foot, or 3.4%.

This prevalence is consistent with that found by the international consensus on diabetic foot, where the global prevalence of diabetic foot was between 3 and 10% (11).

Thus, Sussman et al. found in Great Britain the annual incidence to be between 2 and 3% while the annual prevalence is between 4 and 10% (12).

However, this prevalence is higher than that found by Ramsey et al (13) in the United States, which was 1.9%.

This prevalence could be explained by diabetic patients’ abstention from monitoring on the one hand, but also by their adherence to therapeutic measures and follow-ups.

### Epidemiological data

#### Age and sex

In our study, the mean age was 46.84± 22.4 years with extremes of 3 years and 80 years, and patients over 35 years of age were the most affected and represent 76% of our series.

Age is an influential factor in the occurrence of diabetic foot ulcers, but the study by Awalou MD et al. conducted in Togo found a predominance of the 50+ age group, with a percentage of 40.3% (14). In contrast, the study conducted by Lokrou A and Dago P found that patients with diabetic foot ulcers had an average age of approximately 60 years (15).

This age difference can be explained by the young age of the population in South Kivu, but especially by poor treatment adherence among our patients. The reasons for this poor adherence are numerous: non-acceptance of diabetes, traditional medicine, beliefs, and above all, poverty.

As for sex, in our study, diabetic foot had a female predominance with 68% with a sex ratio of 0.47.

This male predominance was found by Samake D, a female predominance with 61.7% and a rex-ratio of 0.62. On the other hand, Sani et al (16) had found a sex ratio of 2.46.

Poor adherence to treatment, generally recognized in men, would explain this male predominance, but also exposure to trauma, and the frequency of atherosclerotic lesions in men, as well as the importance given by women to care and hygiene.

### Origin

In this study, the majority of patients were from urban areas, representing 64%.

This result is consistent with the results found by El Hariri (2008) (17) with 64% of patients coming from urban areas.

But Dia D et al. found a predominance of patients from the middle class with 58.5%

The urban predominance can be explained by the level of education of the urban population, who undergo check-ups to monitor their illness, while the rural population lacks the means, but also by a lack of information on monitoring their pathology.

### Occupation

In our study, 40% of the patients were professionals

Our results are similar to those found in Senegal by Dia D et al. with a predominance of housewives at 34% of cases(18).

In addition, the urbanization problem in our African societies for the civil servant class has led to changes in diet with excessive consumption of salt and fatty foods and an increase in sedentary lifestyles, exposing our populations to metabolic syndrome.

### Etiologies

#### Family history

In this study, 36% of diabetic patients had a hereditary history of diabetes.

Our results are consistent with those of Kantar Health(19), which assessed the French public’s perception of diabetes and revealed their level of knowledge. In 2012, Kantar Health conducted a survey of a thousand adults, 75% of whom believed that lifestyle was the main risk factor for diabetes, while only 39% (and 22% of diabetics) cited heredity.

This is explained by the genetic predisposition which is often at the basis of type II diabetes, and the lack of voluntary screening of our population for this disease.

### Medical history

In our series, high blood pressure was the most frequent associated disease with 36% followed by obesity in 5 patients or 20%.

This result is identical to that found by Awalou MD et al. (14) with 41.9% of cases of hypertension. However, this result is significantly higher than that found by Monabeka H & Nsakala-Kibangou N at the Brazzaville University Hospital in 2001 (7) with 19% of cases of hypertension and the study by Tadili (17) carried out at the Mohammed VI University Hospital in Marrakech in 2008 with a percentage of 49%.

Dyslipidemia was found in 16% of our patients, which does not agree with the percentage found in Tadili’s study (20), which is 36%. Whereas in EL Allali’s series, this percentage was 7% (21). 20% of our patients were chronic smokers, which agrees with the percentage found in the EL Allali study (21) which is 22% as well as the value found in the Tadili study which is 20%.

In 8% of our cases, the background was not mentioned.

### Clinical data

#### Depending on the type of injury

The most common type of diabetic foot was arteriopathy with 48% followed by neuropathy with 40% and mixed diabetic foot contained 12%.

Gangrene accounts for 48% of diabetic foot lesions in our study, while skin infections account for 32%.

A study conducted in Brazzaville by Monabeka H & Nsakala-Kibangou N gives similar results and shows that arteriopathy was the type of diabetes observed in their study with 52% and gangrene was found in 33.1% of the cases followed(7).

But Amoussou-Guenou et al., in their study, found that the lesions were due to trauma in 32.86% of cases and to burns in 2.86%.

Foot lesions were dominated by gangrene (61.29%) and ischemic necrosis (12.90%)(22).

The predominance of gangrene as a common type of lesion in diabetic foot in our study is explained by its occurrence, which is caused by arterial obstruction due to embolism, shock, infection, or exposure to intense cold. Its origin is most often linked to the prolonged interruption or extreme slowing of blood flow. In past centuries, poorly treated and infected war wounds led to gangrene and then limb amputation. In the absence of oxygen supply, the vessels are damaged, and then the tissues die and putrefy (8).

The occurrence of neuropathy is also explained by the population’s tendency to walk barefoot, especially in rural areas, due to a lack of financial resources and access to manufactured goods. All of this stems from a lack of road connections to cities.

### Depending on the duration of the injury

The consultation time is an important factor that determines the effectiveness and speed of treatment for diabetic foot.

In our study, 36% of patients came for consultation more than a month after the onset of the lesion, hence the extreme of 40 and 365 days, the average number of days for consultation was 26 days. In the study by Awalou MD et al.(14), this delay was less than one month with extremes of 6 and 120 days.

In our series, this is explained by ignorance of the particularities of foot lesions in diabetics as well as the lack of patient education on this problem.

### Depending on the type of diabetes

In our study, type 2 diabetes was the most frequent cause of foot ulcers in diabetic individuals, with a frequency of 68%.

According to the literature review, diabetic foot is common in patients with type 2 diabetes.

This predominance can be explained by the frequency and pathophysiological characteristics of this type of diabetes, including the presence of degenerative pathologies at the time of diagnosis and the long silent progression.

### Patient progression

#### Evolution

In our study, 32% of diabetic patients had amputations while 68% of them received local care. The opposite was observed in 2021, by Dia D et al. Who found an amputation rate of 15.8%(23). The poor glycemic control of our patients confirms the fact that infection in general, and diabetic foot in particular, are factors in the imbalance of diabetes which would eventually lead to necrosis with invasion of part of the lower limb and would lead to amputations.

### Prognosis

In our study, the mortality rate for diabetic foot was 16%.

Incomparable to that of Lokrou which observed a mortality of 6.52% (15).

This mortality is explained by a delay in the care of patients within the care service but also associated with the multiple compilations of diabetes.

## Conclusion

This work entitled **"** *Epidemiology of diabetic foot in Bukavu* " had as its main objective to determine the frequency of diabetic foot at the PANZI General Reference Hospital during the period between January 2017 and December 2021.

We have a retrospective, cross-sectional method with an essentially descriptive aim to achieve our objectives.

During our study, we recorded 745 hospitalized patients, of whom 25 were selected for this study, including:

- The incidence of diabetic foot was 3.4%.
- The mean age was 46.84± 22.4 years with extremes of 3 years and 80 years, and patients over 35 years of age were the most affected and represent 76% of our series.
- A female predominance with 68% of the cases studied with a sex ratio of 0.47.
- Hypertension is the predominant risk factor associated with this condition, accounting for 36%.
- Arteriopathy with 48%.
- Gangrenous lesions represent 48%;
- a predominance of type 2 diabetes, i.e. 68%.
- 94% of patients admitted to the surgical department of the PANZI general referral hospital were discharged with their injuries stabilized.

## Recommendations

To reduce the frequency of diabetic foot, we make the following recommendations:

**For diabetics:**

- Avoid wearing unsuitable footwear;
- Avoid any maneuvers at foot level;
- Consult a doctor immediately if you experience a problem with your lower limb.

**To the general population :**

- Participate in diabetes screenings.

**To the service providers :**

- To perform podiatric examinations at each consultation of a diabetic patient;
- To refer or evacuate the at-risk foot to specialized centers in a timely manner;
- To organize the hearing sessions in the departments.

**To the administrative and political authorities :**

- To establish statistics on diabetic foot across the entire level;
- To provide the HGR/PANZI with a good road communication route.

## Data Availability

All data produced in the course of this study are available from the authors upon reasonable request. All data produced in the course of this work are included in the manuscript.

---

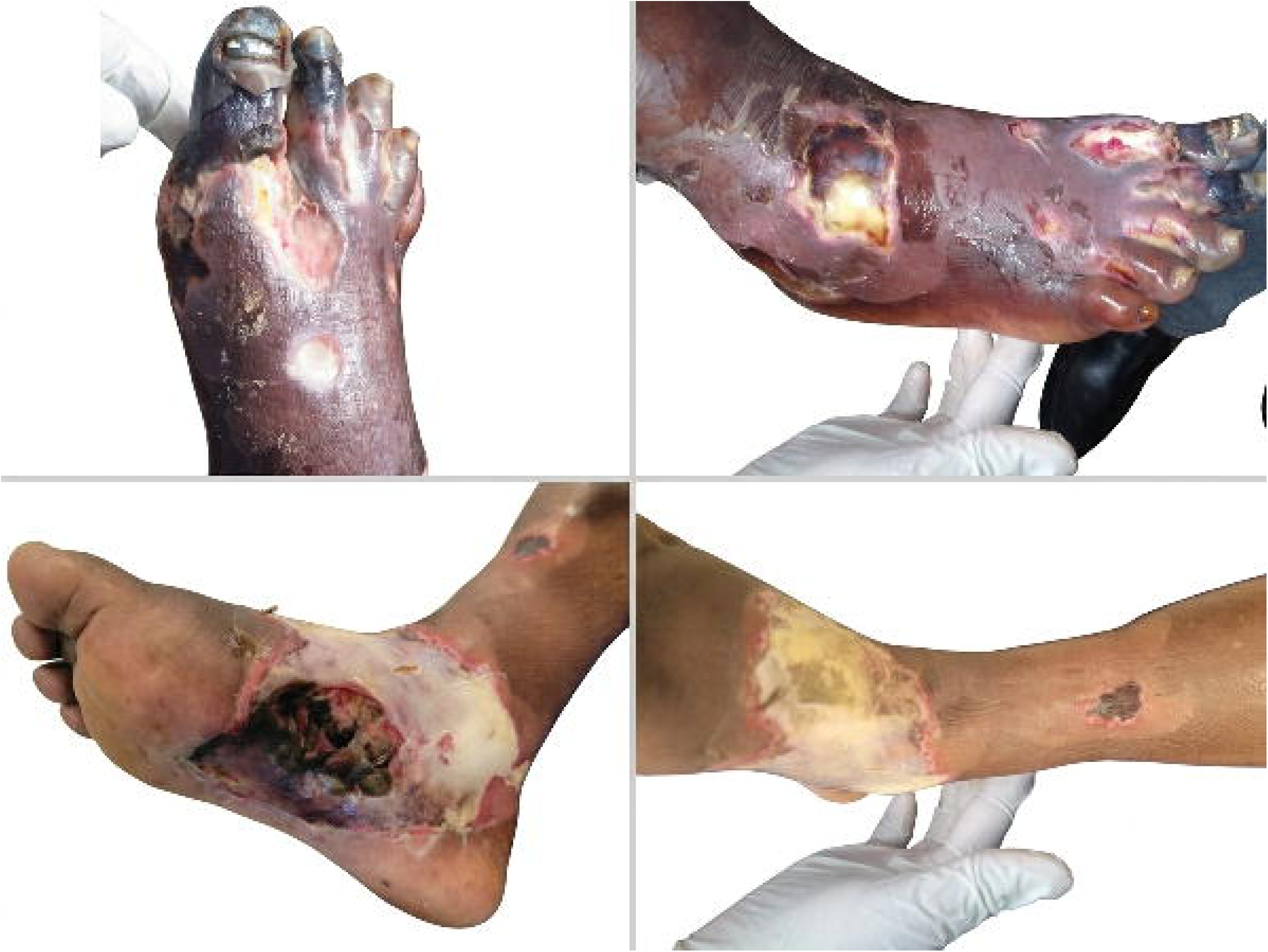

